# Grey or white: what matters? Fraction of white matter tract fibers recruited by deep brain stimulation is causally related to tremor suppression

**DOI:** 10.1101/2023.12.04.23296587

**Authors:** Nanna E.G. Hartong, Matthias Deliano, Jörn Kaufmann, Catherine M. Sweeney-Reed, Jürgen Voges, Imke Galazky, Lars Büntjen

## Abstract

**Background:** Deep brain stimulation (DBS) targets grey matter structures for most clinical indications, such as the thalamic ventral intermediate nucleus (VIM) to treat essential tremor (ET). Alternatively, white matter tracts like the dentatorubrothalamic tract (DRTT) in ET have been suggested to be the actual effector sites of DBS. A direct link between excitation of myelinated fibers and clinically relevant behavior, however, is missing. Here, we retrospectively analyze clinical measurements in patients assessed for VIM-DBS to test the hypothesis that tremor suppression is directly related to the fraction of DRTT-fibers recruited by DBS.

**Methods:** Tremor intensity was accelerometrically quantified at 100 different electrode contacts in 15 patients, while stimulation amplitude was systematically varied. Contact positions were located by stereotactic x-ray imaging. We determined the fraction of fibers recruited within the range of effective DBS-spread by diffusion tensor imaging (DTI) and probabilistic fiber tracking.

**Results:** Utilizing regression analysis, we found that the fraction of activated DRTT-fibers was linearly related to tremor suppression (F(1,592) = 451.55, p < 0.001) with a slope of 1.02 (95% confidence interval [0.93, 1.12]), i.e., relative tremor suppression matched identically the fraction of recruited DRTT-fibers.

**Conclusion:** Our results show that tremor suppression by DBS is causally related to the recruitment of DRTT-fibers and that clinically relevant behavioral effects of DBS can be already predicted from fiber densities pre-operatively. Our analysis approach would enable retrospective identification of DBS effector sites in neuropsychiatric diseases, as well as personalized prospective planning of DBS, substantially reducing intra- and post-operative clinical testing time.

**What is already known on this topic:** Previous studies have demonstrated correlations between clinical outcome in essential tremor suppression by DBS and electrode contact distance to the DRTT. In order to prove that the DRTT is the actual effector site of DBS a direct, a quantitative link between excitation of DRTT fibers and tremor suppression is required.

**What this study adds:** Our study shows that the percent tremor suppression identically matches the fraction of DRTT-fibers recruited by DBS up to a constant offset demonstrating a causal link between tremor suppression and DRTT excitation.

**How this study might affect research, practice or policy:** Our finding solves a long standing dispute and paves the way for novel network interventions through deep brain stimulation. Our analysis approach further paves the way for novel connectomic DBS-targeting strategies. It would allow for personalized prospective planning of DBS substantially reducing intra- and post-operative clinical testing time. It could also be key for the retrospective identification of novel effector sites among candidate sites in various neuropsychiatric diseases.

## Introduction

Deep brain stimulation (DBS) is a well-established treatment for essential tremor (ET) and other movement disorders^1-3^, and has also been successfully applied in various neuropsychiatric pathologies.^4-10^ Grey matter structures are the DBS targets of almost all established clinical indications, with the implicit assumption that the target nuclei constitute the site of DBS action. Thus, the ventral intermediate nucleus of the thalamus (VIM) is considered an effective and safe target in therapy of refractory tremor.^11^ However, white matter tracts containing large myelinated axons with low excitation thresholds must be considered as potential effector sites of DBS, as well.^12,13^ In a connectivity-associated segmentation of the thalamus in tremor patients, VIM-DBS was most effective when the contact position with the highest connectivity to the dentate nucleus was chosen.^14^ The anatomical basis of this connectivity is the dentatorubrothalamic tract (DRTT).^15^ Using deterministic fiber tracking based on diffusion tensor imaging (DTI), Coenen *et al*. showed that electrode contacts associated with greater therapeutic effects have mostly been localized in the immediate vicinity of the DRTT.^16^ Dembek *et al*. reported a correlation between electrode contact distances to the DRTT and the current required for tremor control.^17^ This led to a debate about whether the VIM or the DRTT is the actual DBS effector site in ET suppression.^18^ The idea of a white matter effector site has been transferred to the treatment of other movement disorders^19^, and the term “connectomic DBS” has been coined to encapsulate the idea of white matter DBS into a coherent concept for better understanding the network effects of DBS.^20^

To date, evidence for connectomic DBS effects has been based on indirect correlations between clinical rating scales and contact distances to fiber tracts. A direct, quantitative and causal relationship between excitation of tract fibers by DBS and objectively measured behavior in individual patients has not yet been demonstrated. Here, we test the hypothesis that tremor suppression is directly and quantitatively linked to the fraction of DRTT-fibers excited by DBS.

## Materials and methods

### Patients and study design

We report an observational study retrospectively analyzing clinical routine measurements in patients assessed for VIM-DBS. Patients were implanted with DBS electrodes at the University Hospital Magdeburg between 09/2012 and 12/2017. The data consisted of quantitative tremor measurements and diffusion tensor images (see below). Our study is part of a larger study starting 03/2018 investigating the influence of tremor intensity and neuromodulation by DBS on motor learning. It is registered at the German Clinical Trial Register (DRKS00032906), is conducted in accordance with the guidelines of the Declaration of Helsinki and approved by the Ethics Committee of the Medical Faculty (Approval no. 25/18). Inclusion criteria were age >18 years, diagnosis of ET for at least five years, and 12 months minimum follow-up. Exclusion criteria were neurological conditions and comorbidities limiting participation (e.g., dementia), and tremor-inducing movement disorders other than ET. Fifteen patients (six male, nine female) met the inclusion criteria and agreed to participate. Their mean age at surgery was 70.5+/-10 years (median: 72 years), the mean duration of illness in this patient group was 22.7+/-14.2 years (median: 16 years) before operative treatment. Notably, the patient group had already received electrodes implanted for DBS before the hypothesis addressed here had been posed. Also, the clinical examiners and the patients were blinded to the electrode location with respect to VIM and DRTT during tremor measurement, as the actual locations were determined only after data collection.

### Imaging and probabilistic fiber tractography

Patients received preoperative T1-, T2-, proton density, and diffusion-weighted MRI-scanning. A 3-Tesla scanner (Siemens Magnetom Verio 3T, 32-channel head coil, T1-sequence: 3D-magnetization prepared rapid acquisition with gradient echoes^21^, DTI: 30 non-collinear diffusion directions, b = 1000 s/mm^2^) was used in 13 patients and a 1.5-Tesla scanner (Philips Intera 1.5T, 16-channel neurovascular head coil, T1-sequence: gadolinium-based contrast, DTI: 15-direction gradient scheme on average, b = 1000 s/mm^2^) in two patients. We applied patient-specific probabilistic fiber tracking with a two-stage Monte Carlo algorithm^22^. We first manually segmented three regions of interest (ROI): primary motor cortex in the T1-images, and ipsilateral red nucleus and contralateral dentate nucleus in the T2-images.^15^ DRTT-fibers were then identified by starting in primary motor cortex as seed region (precentral gyrus, with 100,000 starts per voxel) and crossing the ipsilateral red nucleus to reach the contralateral dentate nucleus (filter regions). Density maps were created representing the frequency of fibers crossing the relevant voxels (Fig. 1A) and transformed into the T1-matrix.

**Figure 1.**
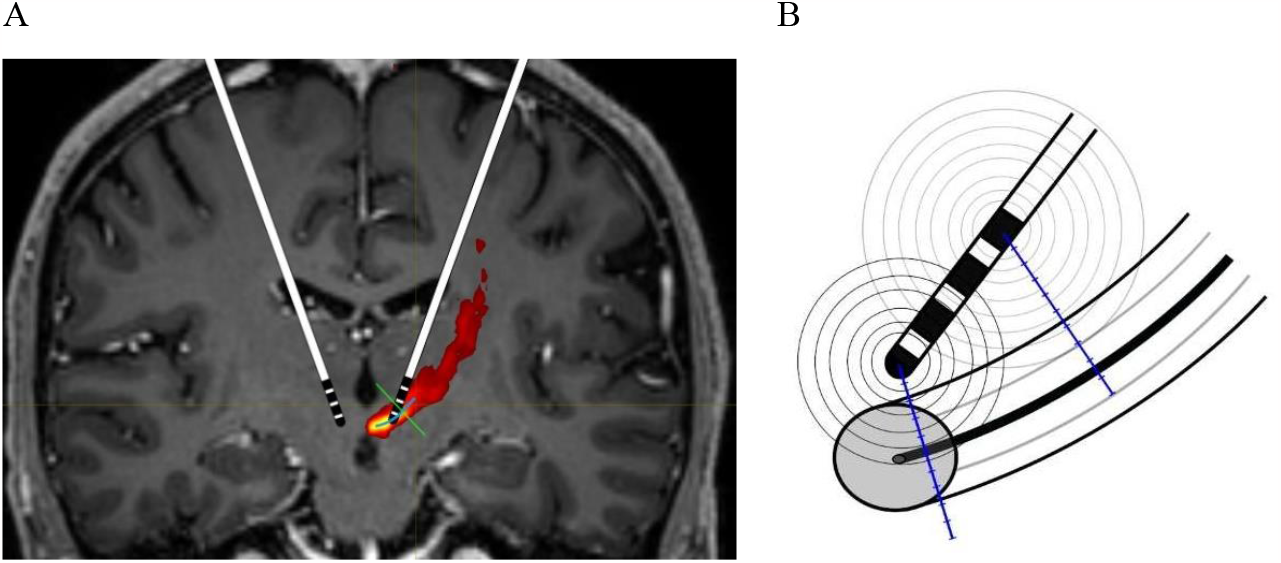
Spatial relationships between implanted DBS-electrode and DRTT. (**A**) Illustration of DBS-electrode implantation relative to the DRTT. The fiber density map is color-coded from red (low density) to yellow (high density). The DBS electrode is schematically drawn along its planned trajectory and at its planned target coordinates. (**B**) Schematic depiction of spatial relationships between electrode contacts, effective DBS-spread (black rings), and the DRTT. Analyses were carried out along the shortest projection line from a given contact through the DRTT, perpendicular to the trajectory of the center of gravity of the DRTT (DRTT-COG).

Center points of the four electrode contacts or segmented electrode-rings were determined from intraoperative stereotactic x-ray images in the anterior–posterior and lateral plane co-registered with the preoperative MRI images and a postoperative stereotactic CT. Stereotactic coordinates (Fig. 2A) were determined in relation to the posterior commissural point (PCP).

**Figure 2.**
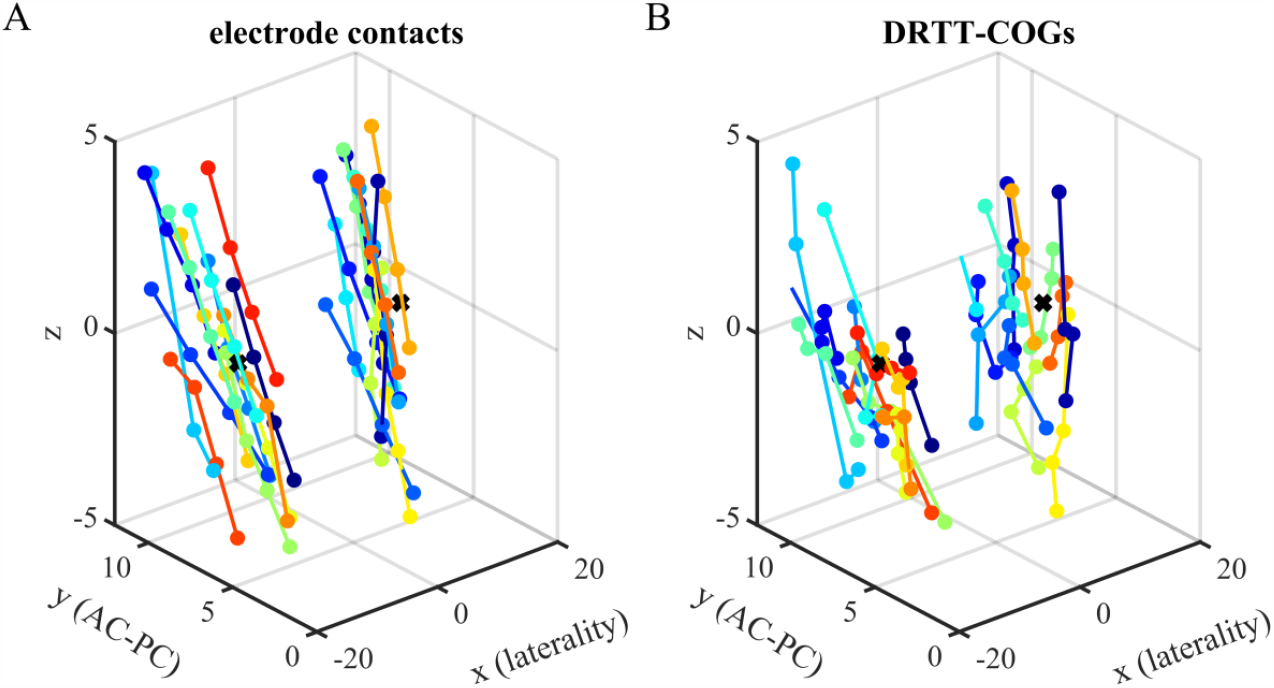
Measured positions of electrode contacts and nearest DRTT centers of gravity. (**A**) Positions of electrode contacts were extracted from intraoperative stereotactic x-ray imaging in each patient. The four electrode contacts (dots) are connected by a line. The line color indicates the electrode. The x-axis indicates laterality, the y-axis is oriented along the AC-PC line. The VIM atlas coordinates are indicated by a black cross. (**B**) Positions of DRTT-COGs obtained from fiber density maps by DTI-based probabilistic tractography in the same coordinate system as the contacts in A.

### Planning & surgical procedure

The target point in the VIM was chosen according to atlas coordinates in relation to the PCP (x: +/- 13.5 mm, y: +7 mm, z: 0 mm, Schaltenbrand Wahren Brain Atlas^23^) in T1-images. We corrected for patient-specific distortions along the line between the anterior and posterior commissure (AC-PC). Trajectories were routinely prolonged by four mm to position at least one contact in the zona incerta. The surgeon determined the final target location in each patient individually, considering fiber tracking information, vessel configurations and intraoperative neurological testing. Electrodes were implanted under local anesthesia. After finding the optimal implantation depth via macrostimulation during awake surgery, patients received one of the two following hardware configurations: Medtronic 3389, Activa PC (Medtronic Inc., Minnesota, USA), or St. Jude Medical Directional Lead/Infinity (St. Jude Medical, Minnesota, USA; from 2017 Abbott Laboratories, Illinois, USA). Implantable pulse generators (IPGs) were placed subcutaneously under general anesthesia.

### Tremor analysis

Tremor accelerometry^24,25^ was carried out with the triaxial wireless accelerometer ACL 500 (Biometrics Ltd., Newport, UK) prior to the study for diagnostic purposes. Ten patients underwent bilateral measurement, from one electrode in each hemisphere. Five patients displayed only one-sided visible tremor, and therefore received only unilateral measurement. Following a standardized testing protocol for each hemisphere, IPGs were switched off for at least 12 hours before testing. Patients paused tremor-suppressing drugs on the day, and nicotine and caffeine at least two hours before measurement. Temporary and persisting side-effects were monitored. In case of intolerable paresthesia persisting for over 60 s, testing was terminated. Each hemisphere was tested separately applying DBS at a frequency of 130 Hz and 60 μs pulse-width, while the contralateral electrode was switched off. Segmented contacts were stimulated in ring-mode. Patients sat upright with the accelerometer attached to the intermediate phalanx of the middle finger. Patients were asked to hold their arms horizontally in front of them at shoulder height, palms upwards, for ten seconds. Three consecutive measurements were first performed without DBS (baseline tremor) and followed by increasing amplitude in 0.5 mA (Abbott) or 0.5 V (Medtronic) increments. After each measurement, there was a 30 s break, and at least a two hour pause before switching hemispheres.

Tremor intensity was determined as magnitude of the acceleration vectors (three orthogonal sensor axes) obtained by the DataLITE software (version 10.05, Biometrics Ltd., Newport, UK). Intensity values were averaged from one second after starting to one second before completing the arm-holding task. Tremor intensities measured at different DBS-amplitudes and contacts were subtracted from the average of the three baseline values as a raw measure of tremor suppression. A robust measure of DBS-related relative tremor suppression was calculated as a percentage by subtracting the minimum tremor suppression across the four contacts of an electrode, dividing by the maximum across contacts, and multiplying by one hundred.

### Effective spread of activation from electrode contact to DRTT fibers

We assumed that DRTT fiber recruitment is largely determined by the radial spread of DBS through the DRTT^12,13^ which we determined for each contact and DBS-amplitude from a well-established amplitude–distance relationship of electrical brain stimulation^26^ adapted to voltage-driven DBS^27^:

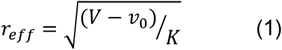

*r*_*eff*_ is the radius of effective neural activation, *V* the staimulation voltage, *K* a voltage-distance constant, and *v*_*0*_ threshold offset. For current stimulation, amplitudes were transformed into voltage using Ohm’s law assuming a tissue resistance of 1.0 kΩ.

### Combined DRTT-centered analysis of fiber density and tremor suppression

We reconstructed DRTT fiber density profiles radially through the DRTT, along the shortest projection line from the contact, i.e., perpendicular to the trajectory of the DRTT-center of gravity (DRTT-COG) (Figs. 1B and 2B). Tractographic density values were determined in steps of 0.5 mm along this line. To relate tremor suppression to DRTT fiber recruitment, we spatially aligned our contact-centered measures to the DRTT-COG by subtracting the distance between each contact and the DRTT-COG on the projection line. To increase the range and spatial resolution, we pooled the values of effective DBS-spread, its corresponding relative tremor suppression, DRTT-densities, and recruited DRTT fractions across the four contacts of an electrode. We sorted the values by effective DBS-spread and distance relative to the DRTT-COG and smoothed them.

### Statistical analysis

For statistical analysis, we employed mixed regression models implemented in the nlme-package of R (https://www.r-project.org). Percentage values were normalized by a logit transformation. As we expected tremor measurements to be correlated, regression models included random effects grouped by the factors *Patient, Side* and *Contact*. Models were corrected for serial correlations, and variance inhomogeneities.

In the contact-centered frame, we analyzed the dependency between relative tremor suppression in percent as response variable and the treatment variables – effective DBS-spread (*Spread*) and contact distance to the DRTT-COG (*Dist_DRTT*) and the VIM (*Dist_VIM*) – and non-specific factors, including stimulation mode (*Mode*), stimulation frequency (*Rate*), implantation side (*Side*), and sex (*Sex*). We hypothesized that for a brain structure to be a DBS effector site, there must be an interaction between *Spread* and the contact distance to this structure (*Dist_DRTT* or *Dist_VIM)*.

In the DRTT-centered frame, we statistically analyzed the relationship between relative tremor suppression as response variable in percent and the fraction of recruited DRTT fibers in precent within the range of effective DBS-spread (*Fract*) for the pooled, sorted, and smoothed values aligned relative to the DRTT-COG.

Eight stimulation amplitudes were applied to four contacts of 25 electrodes, 10 bilaterally, 5 unilaterally. From the possible 800 combinations of contacts and DBS-amplitudes, 711 data points were actually measured according to protocol without side-effects. Data from contacts with negative correlation between stimulation amplitude and tremor suppression (< -0.1) were excluded from further analysis, as they indicate strong influence of side-effects. Data points with residuals exceeding four standard deviations, were excluded as outliers. In total, 597 of the 711 measured data points were used for regression analysis (84%).

## Results

### Contact-centered analysis

DBS-effects depend on both the effective DBS-spread and the distance between electrode contact and effector site. Effective DBS-spread^26,27^ grew nonlinearly with stimulation amplitude (Fig. 3A), affecting neural elements up to four mm around a contact with the range of amplitudes employed. Fig. 2A shows the positions of the electrode contacts and Fig. 2B the corresponding positions of the nearest DRTT-COGs. Contacts were distributed both medially and laterally relative to the atlas position of the VIM center (black cross). DRTT-COGs were located more ventrally than the contacts relative to the VIM. Contacts were located between 0.0 and 6.0 mm away from the DRTT-COG, with a broad peak around 2.0 mm (Fig. 3B, red bars). With respect to the patient-specific VIM target points, contacts were positioned at distances between 0.0 and 8.0 mm with a peak between 3.0 and 4.0 mm (blue bars).

**Figure 3.**
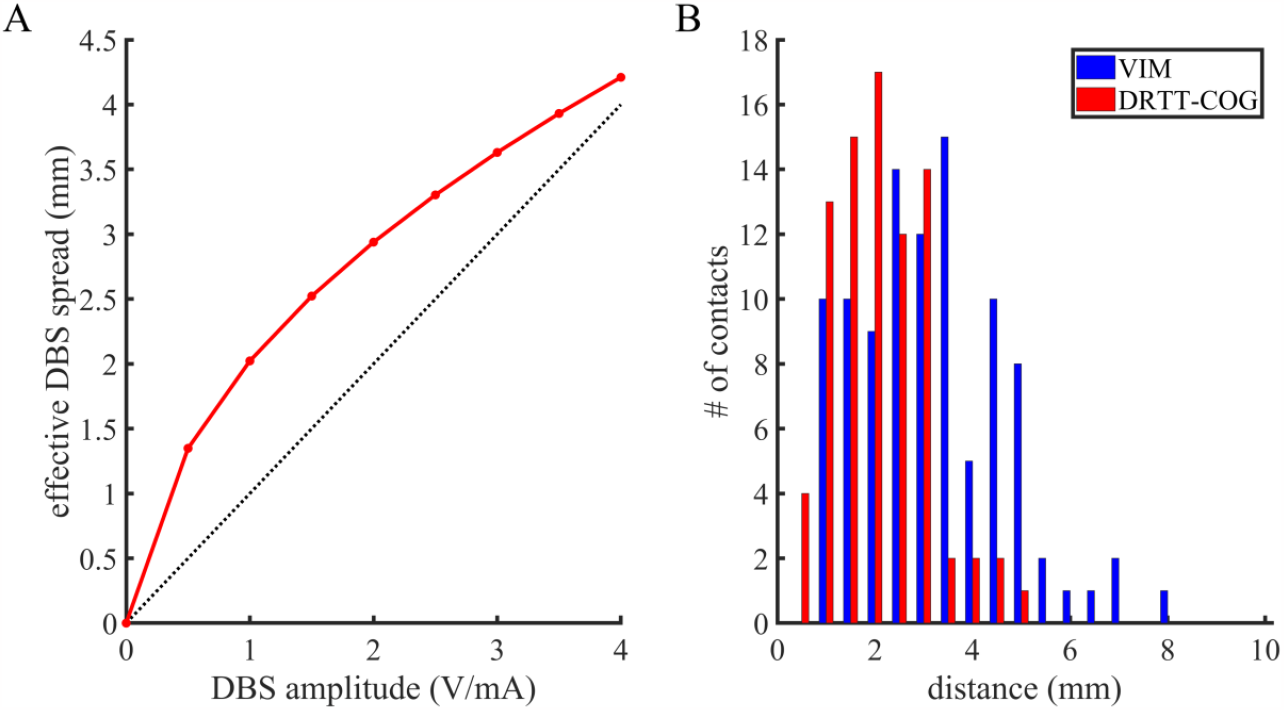
Effective DBS-spread and contact distances to the DRTT and the VIM. (**A**) Effective DBS-spread obtained from a simple amplitude–distance relationship, adapted to DBS by Kuncel et al.,^27^ based on a distance constant K (red curve), determined separately for voltage and current modes of DBS. (**B**) Distribution of patient-specific anatomical distances between electrode contacts and the corresponding DRTT-COGs (red bars), or the individual VIM target points corrected for patient-specific distortions along the AC-PC line (blue bars) in mm. Distances to the DRTT were measured along the shortest perpendicular projection line onto the DRTT-COG trajectory.

Regression analysis revealed a highly significant main effect of the DBS-spread from the contact (*Spread*: *F*(1,511) = 430.14, *p* < 0.0001), and a significant interaction between effective DBS-spread and contact distance to the DRTT-COG (*Spread* x *Dist_DRTT*: *F*(1,511) = 4.43, *p* < 0.05), but no interaction between spread and distance to the VIM (*Spread* x *Dist_VIM*: *F*(1,511) = 1.83, *p* > 0.18). The interaction between spread and distance to the DRTT-COG is illustrated by the grand mean in Fig. 4. For electrode contacts with distances less than 3.0 mm from the DRTT-COG (Fig. 4A, red curve), higher tremor suppression was found than for more distant contacts (blue curve), but only at mid-level stimulation. No difference in tremor suppression was observed between contacts near and distant to the VIM (Fig. 4B). In a control analysis, we did not find effects of non-specific factors (*Sex, Mode* and *Side*: F(1,9) < 0.21, *p* > 0.65, and *Rate*: *F*(3,9) = 0.40, *p* > 0.75).

**Figure 4.**
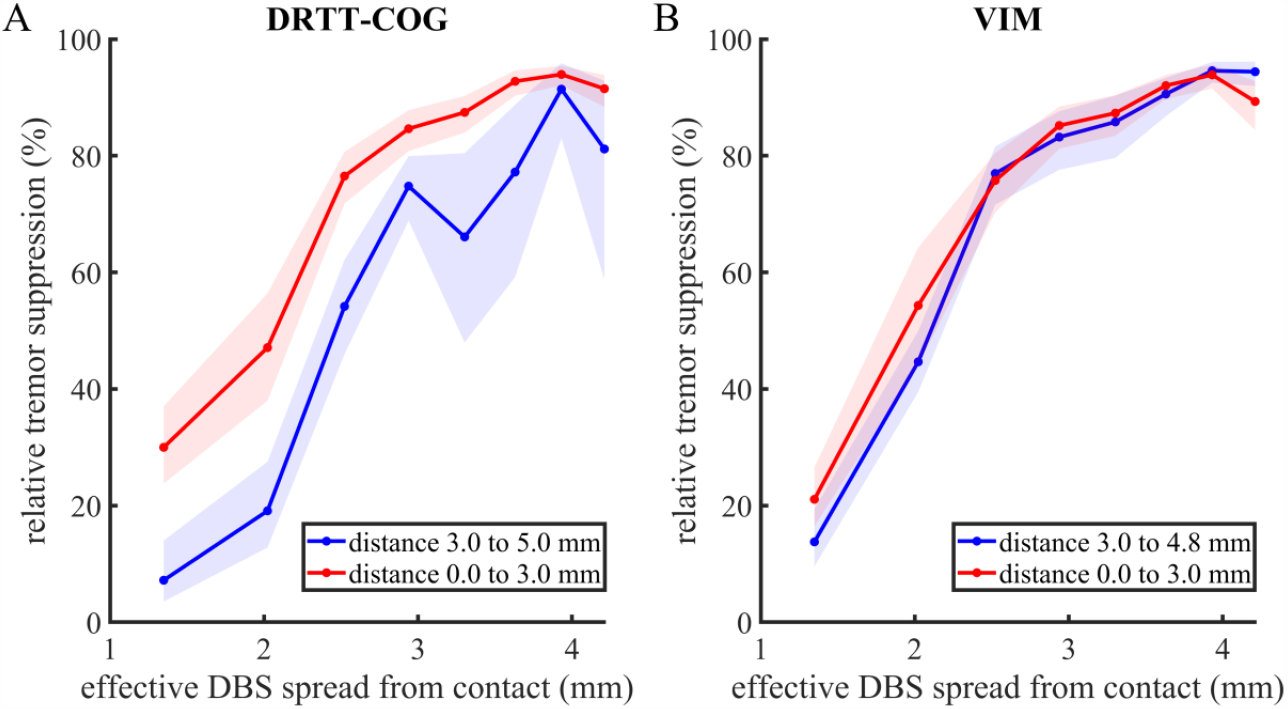
Relative tremor suppression as function of effective DBS-spread and contact distance to DRTT. (**A**) Relative tremor suppression in percent as a function of effective DBS-spread, determined from the amplitude–distance relationship shown in Fig. 3A. The curves show the grand mean of relative tremor suppression averaged across contacts near the DRTT-COG (red curve) less than 3.0 mm away from it, and more distant contacts (blue curve) equal or more than 3.0 mm away from the DRTT-COG. Shaded areas indicate the standard error of the mean (SEM). (**B**) Relative tremor suppression as in A, but for the electrode contacts near the VIM (red curve), less than 3.0 mm away from it, and more distant contacts (blue curve), more than 3.0 mm away from it.

### DRTT-centered analysis

To test our hypothesis that the degree of tremor suppression is determined by the fraction of DRTT fibers recruited by DBS, we spatially aligned our measures to the DRTT-COG. Fig. 5 shows the density profiles of DRTT fibers in distance relative to the DRTT-COG along the shortest projection line from each contact to the DRTT (Fig. 1B). In the grand mean across patients and hemispheres, measured DRTT-densities (Fig. 5, blue curve) fitted well to a Gaussian model with variable mean, standard deviation, amplitude, and offset for each contact. The red curve in Fig. 5 shows the grand mean of the Gaussian curves obtained from model fits for each contact (red curve), and for the median of the parameters across contacts (dashed black curve). For ten out of the 100 contacts, fitting did not converge, and the median values of the parameters of the neighboring contacts were used. The maximum of the measured profiles and the fitted curves were all found at the DRTT-COG (mean value). The grand mean half-width at half-height was 6.29 +/-0.45 mm, as obtained from the standard deviations of the Gaussian model fitted to each contact.

**Figure 5.**
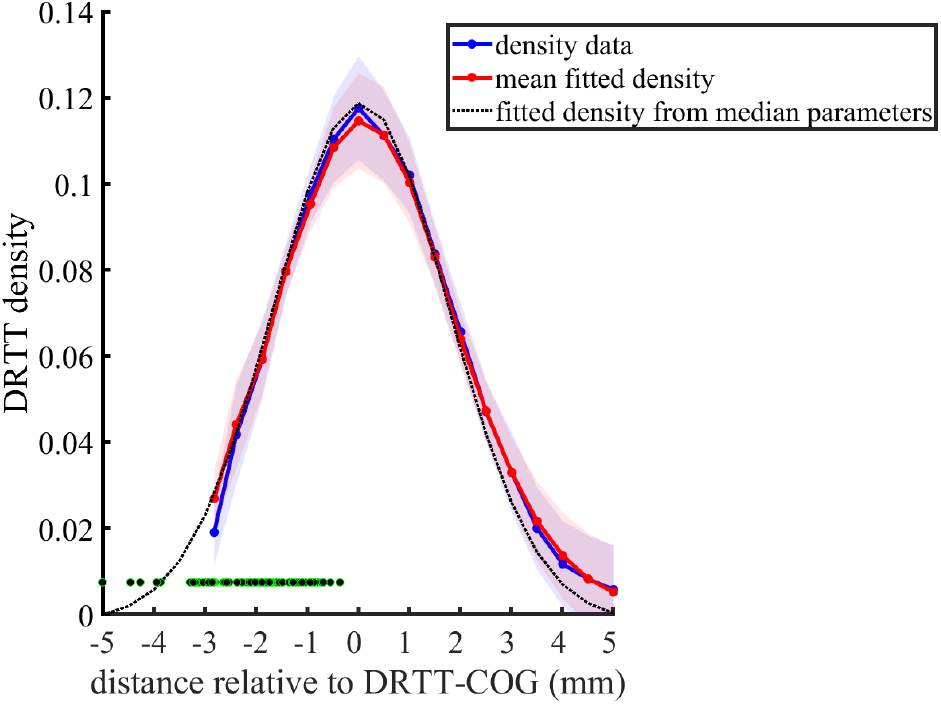
Profile of DRTT fiber density in the vicinity of the electrode contacts. DRTT fiber density as a function of the distance relative to the DRTT-COG. DRTT density profiles were obtained from DTI-based probabilistic fiber tracking in steps of 0.5 mm along the shortest projection line between each contact and the DRT-COG. The blue curve shows the grand mean density profile across patients and hemispheres (n = 25), smoothed by a sliding window with a half-width of one point, and spatially aligned to the DRTT-COG. The grand mean was calculated across electrodes from the median across the four electrode contacts. The red curve displays the grand mean of the continuous Gaussian model curves fitted to the aligned tractographic DRTT density values for each contact, and the dashed black curve shows the Gaussian model curve obtained from the median of the fitted parameters across all contacts and electrodes. Shaded areas indicate the 95% confidence interval. All density profiles were normalized to their integrals (sums). Electrode contact positions relative to the DRTT-COG are marked with green dots.

From the fitted mean and standard deviation of the Gaussian density profiles, we calculated the fraction of DRTT fibers activated within a radius of effective DBS-spread relative to the DRTT-COG via a cumulative Gaussian distribution. To increase spatial resolution, values of tremor suppression and recruited DRTT fractions were pooled across contacts and sorted by effective spread. After removing an observed constant offset by subtracting the mean difference between the percentage of tremor suppression and the percentage of activated DRTT fibers across the whole range of effective spread, the increase in relative tremor suppression identically matched the increasing fraction of recruited DRTT with increasing effective DBS-spread (Fig. 6A). A mixed regression model corrected for spatial correlations possibly introduced by the smoothing (Fig. 6B) showed a highly significant effect of the recruited fraction of the DRTT on tremor suppression (*F*(1,592) = 451.55, *p* < 0.001). The estimated slope was 1.02, with a 95% confidence interval ranging from 0.93 to 1.12. By removing the offset, the intercept of -0.01 was close to zero. Its 95% confidence interval ranged from -0.18 to 0.17, including zero, and a regression coefficient not significantly different from zero (*F*(1,592) = 0.30, *p* > 0.58). Therefore, relative tremor suppression identically matched the fraction of activated DRTT fibers up to a subtracted constant.

**Figure 6.**
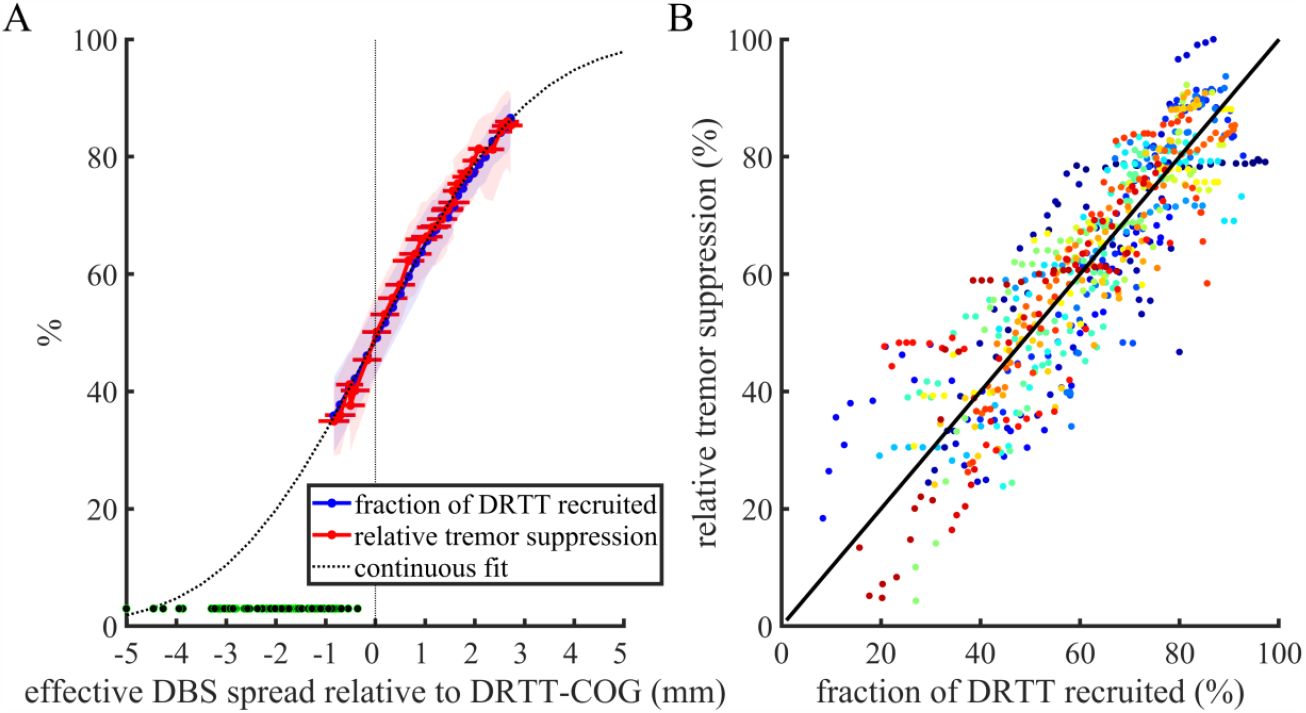
Fractions of recruited DRTT fibers and relative tremor suppression as function of effective DBS-spread relative to the DRTT. (**A**) Grand means of relative tremor suppression (red curve), and fraction of DRTT fibers activated (blue curve) as a percentage. DRTT fractions were determined from the parameters of the Gaussian model fitted to tractographic density values (see Fig. 5), as a function of the effective DBS-spread relative to the DRTT-COG. All values were pooled across contacts, sorted by effective DBS-spread, and smoothed with a bandwidth of three data points. A constant offset was subtracted from the relative tremor suppression. Shaded areas indicate the 95% confidence interval of tremor suppression and activated DRTT fractions. Red horizontal error bars display the 95% confidence interval of the effective DBS-spread. Electrode contact positions relative to the DRTT-COG are marked by green dots. (**B**) Relative tremor suppression in relation to the fraction of recruited DRTT fibers for each single electrode. Colors distinguish data from electrodes implanted in different patients and on different sides within a patient.

## Discussion

Relative tremor suppression significantly depended on an interaction between effective DBS-spread and contact distance to the DRTT. Tremor suppression was stronger at lower DBS-amplitudes when the contact distance to the DRTT was smaller. No interaction effect between spread and contact distance was found for the VIM. Such a combined effect of effective DBS-spread and contact distance is a necessary precondition for a brain structure to be an effector site of DBS, which was the case for the DRTT, but not for the VIM.

However, significant correlations of effective DBS-spread and contact distance to a brain structure per se are not sufficient for the structure to be a DBS effector site. Stimulated brain structures spatially correlated with the effector site might still explain this effect. We therefore carried out a direct analysis of the effect of DRTT stimulation on relative tremor suppression based on its fiber densities. After spatially aligning our measures relative to the DRTT-COG, the fraction of DBS-activated DRTT fibers had a highly significant effect on relative tremor suppression. Moreover, the CI of the slope between tremor suppression and the fraction of recruited fibers included one. Therefore, tremor suppression matched identically the fraction of recruited DRTT-fibers up to a constant offset. Notably, no other factors not directly related to our hypothesis, such as sex, DBS mode, and implantation side, affected tremor suppression. Together, these findings therefore provide strong evidence for a quantitative causal relationship between the fraction of recruited DRTT fibers and relative tremor suppression.

While the VIM is a well-established target for DBS to treat pharmacoresistant tremor, some group analyses have already suggested that the DRTT could be the key structure in DBS for tremor control.^14,16,17^ However, none of these studies demonstrated a causal relationship between DRTT fiber excitation. One main strength of our study is the precise accelerometric tremor measurement. In most of the other studies, tremor scores such as the Fahn-Tolosa-Marin tremor rating scale (TRS) were used to rate tremor severity^28^, usually after extensive optimization of stimulation parameters. Rater-independent tremor accelerometry is more objective than these scores in terms of tremor intensity,^29^ although it still displays fluctuations over time.^30^ Moreover, we employed patient-specific probabilistic fiber tracking. Although computationally more laborious than deterministic tracking, it enabled detection of crossing fibers and individual reconstruction of radial fiber density profiles without referring to an “average DRTT” derived from a healthy participant group.^31^

### Grey versus white: network effects leading to tremor suppression

There is growing evidence that movement disorders such as Parkinson’s disease and ET should be considered as so-called circuitopathies^32^ and that tremor is due to a dysfunctional oscillatory network state. In these circuitopathies, fiber tracts like the DRTT function as information highways, a concept for which the term “connectomic DBS” was coined.^20^ The results of our study convincingly underline the validity of this idea for ET. Still, the complete causal chain from local DBS effects to a more global functional network state dampening the tremor remains to be elucidated.^33^ Direct excitation of myelinated axons by DBS is biophysically well understood. High frequency stimulation presumably leads to excitation of DRTT axons. Local suppressing effects, e.g., by changes in the local ionic environment or by membrane changes, might also play a role.^34^ However, the effect of DBS is mediated transsynaptically^33,34^ modulating the oscillatory dynamics of the network connected through the DRTT. Still, our results solve a long standing dispute about whether the DRTT or the VIM is the effector site of DBS in essential tremor treatment. Showing that DBS causes tremor suppression by an activation of axonal fibers of the DRTT connecting nodes of a motor network, and not by stimulating a single network node like the VIM provides a strong foundation for the connectomic DBS concept.

### Towards a patient-specific connectomic targeting strategy based on probabilistic fiber density tractography

Clinical outcome of DBS in suitably chosen patients depends mainly on optimal lead positioning and adjustment of stimulation parameters.^35^ While traditional targeting strategies relied on landmarks and group results, modern imaging technologies allow for individually tailored surgical approaches optimizing treatment results. Grey matter structures like the VIM are often difficult to locate using non-invasive brain imaging,^36^ whereas the anatomy of fiber tracts like the DRTT can be visualized using DTI in individual patients. Information on fiber anatomy can be determined from preoperative MRI, and our findings suggest that DBS planning based on individual fiber density maps is useful in optimizing clinical outcome. Fenoy *et al*.^37^ compared TRS and stimulation parameters after optimization in a 12-month follow-up between a group with indirect, atlas-based VIM-DBS and a group with direct DRTT-DBS using deterministic fiber tracking. Significant tremor suppression was achieved in both groups, with the DRTT group showing lower average voltage, frequency, and pulse width after optimization. While these findings suggest that DRTT-targeted DBS might be more effective than VIM-targeted DBS, our study adds two vital factors that enable evaluation of causality: our stimulation parameter programming was blinded to the target, and we examined the exact location of individual electrode contacts relative to the DRTT.

Our findings and analysis approach could serve as a basis for developing novel connectomic DBS targeting strategies. Patient-specific tractography can be used in prospective DBS planning to optimize contact positions together with DBS-amplitudes for known effector sites. This approach would substantially reduce time for clinical testing and DBS-programming. Our approach also allows retrospective identification of DBS effector sites. Best matches between individual fiber density profiles and treatment effects can be searched among candidate fiber tracts^38^ obtained from functional connectivity analyses and lesion network mapping^39^ by regression model comparisons.

In summary, our findings demonstrate that the DRTT is indeed the anatomical structure causally responsible for therapeutic effects of DBS in ET patients. If our findings are generalizable to other DBS targets and conditions, DTI-based probabilistic fiber tracking will become the new gold standard for pre-operative targeting and for the identification of DBS effector sites, which makes individually tailored personalized DBS possible for all patients.

## Data Availability

Raw data were generated at the University Hospital Magdeburg. Derived data supporting the findings of this study are available from the corresponding author on request.

## Abbreviations

AC-PC: anterior commissure - posterior commissure
CI: confidence interval
CT: computer tomography
COG: center of gravity
DBS: deep brain stimulation
DRTT: dentatorubrothalamic tract
DTI: diffusion tensor imaging
ET: essential tremor
IPG: implantable pulse generator
PCP: posterior commissural point
SEM: standard error of the mean
TRS: tremor rating scale
VIM: ventral intermediate nucleus

## Notes

### Competing Interest Statement

J. Voges occasionally received honoraria from Boston Scientific. There are no other conflicts of interest to declare.
L. Buentjen occasionally has given educational talks for Boston Scientific, Medtronic, SJM/ Abott in the past, but none of them during the last 3 years.

### Clinical Trial

DRKS00032906

### Funding Statement

This research received no external funding. CMSR received personal funding from the Deutsche Forschungsgemeinschaft (SW 214/2-1).

### Author Declarations

The study was conducted in accordance with the guidelines of the Declaration of Helsinki and approved by the Ethics Committee of the Medical Faculty of the Otto-von-Guericke-University Magdeburg (Approval no. 25/18).

